# Feasibility of following a fasting-mimicking diet programme in persons with type 2 diabetes – a mixed-methods study

**DOI:** 10.1101/2025.10.30.25339142

**Authors:** Marjolein P. Schoonakker, Elske L. van den Burg, Carlijn A. Sturm, Hildo J. Lamb, Hanno Pijl, Mattijs E. Numans, Marieke A. Adriaanse, Petra G. van Peet

## Abstract

**Background & Aims:** A fasting-mimicking programme in primary care demonstrated metabolic benefits for persons with type 2 diabetes (T2D). This study evaluated the feasibility of this approach.

**Methods:** Persons with T2D who used metformin or no medication for glycemic control, were randomly assigned to monthly 5-day fasting-mimicking diet (FMD) cycles plus usual care (n=51) or usual care alone (n=49) for one year. In this mixed method study, feasibility of following an FMD was evaluated by combining quantitative data, including reasons for discontinuation, serum ketone levels, and treatment satisfaction, and qualitative data from focus group discussions.

**Results:** In the FMD group, 61% of the participants completed the FMD programme, 31% discontinued the programme due to diet-related issues, and 8% discontinued the programme for other reasons.

Ketone levels were consistently higher in the FMD group than in the control group (p<0.01). Treatment satisfaction did not differ between the groups. Focus groups (n=20) revealed facilitators of adherence, including convenience, short FMD cycles, not feeling hungry, internal motivation, believing in beneficial effects, experiencing health effects and social support. Barriers included taste, quantity, and frequency of the FMD, environmental temptations, and lack of social support.

**Conclusions:** The implementation of periodic FMD cycles appears feasible for T2D patients in primary care, supported by adherence rates, ketone levels, and treatment satisfaction. Adherence might be enhanced by addressing identified facilitators and barriers.

**Trial registration:** ClinicalTrials.gov; NCT03811587. Registered 22 January 2019.

## Introduction

Type 2 diabetes (T2D) significantly impacts both health and healthcare costs, necessitating effective treatments(1, 2). T2D arises from a combination of genetic and environmental factors, with lifestyle being one of the most critical contributors. Lifestyle changes are therefore essential and can even reverse T2D; however, many current treatment options are difficult to sustain(3–9). This underscores the need to expand the range of treatment options to improve adherence by allowing persons to choose treatments that align with their needs and capabilities. Consequently, intermittent and periodic fasting programs have been explored as potential treatment strategies for T2D(8, 10–12).

Intermittent and periodic fasting can positively influence glucose regulation and can lead to a reduction in glucose-lowering medication use in persons with T2D(10–12). Compared with total fasting, fasting-mimicking diets offer similar health benefits(13, 14) and might be more feasible because of the usually limited fasting period of 5 days a month and the availability of light meals during fasting periods(15). As feasibility is a crucial determinant of the long-term effectiveness of a lifestyle intervention, the feasibility of implementing an FMD in individuals with T2D was investigated in the Fasting In diabetes Treatment (FIT) trial(16, 17). This involved evaluating self-reported adherence to the FMD, analysing ketone levels to objectively assess adherence, measuring treatment satisfaction and exploring facilitators of and barriers to adherence to the FMD in focus groups with participants allocated to the FMD group.

## Materials & Methods

### Study design

The FIT trial was conducted at the Leiden University Medical Centre (LUMC) in the Netherlands between November 2018 and August 2021. The FIT trial was a randomized, controlled, assessor-blinded intervention trial(16). The protocol and amendments were approved by the Medical Research Ethics Committee of the LUMC. The trial was registered at ClinicalTrials.gov (NCT03811587) and performed according to the principles of the Declaration of Helsinki, following the Medical Research Involving Human Subjects Act (WMO) and the standards of Good Clinical Practice (GCP). All participants provided written informed consent to participate.

Demographics were collected for participants included in the FIT trial. To assess the feasibility of the FMD program, we employed a convergent parallel mixed-methods design that integrates quantitative and qualitative data(18). Statistical analyses were performed via RStudio version 4.1.0 for Windows. Figures were created in GraphPad Prism version 9.0.1 for Windows and Microsoft PowerPoint version 2308 for Windows.

### Quantitative data

#### Participants and Procedure

Participants were eligible for the FIT trial if they were diagnosed with T2D, had a BMI ≥ 27 kg/m2, were aged between 18-75 years, and were treated in primary care with lifestyle advice only in combination with a glycated haemoglobin (HbA1c) above 48 mmol/mol (6.5%), or lifestyle advice plus metformin as the sole glucose-lowering drug irrespective of HbA1c levels. The participants were randomly assigned to the FMD group, receiving twelve monthly cycles of a five-day FMD alongside usual care, or to the control group receiving usual care only. The FMD comprised complete meal replacement products, with plant-based ingredients (**Appendix 1**). Caloric content and macronutrient composition were as follows; day 1 contained ∼ 4600 kJ (1100 kcal, 10% protein, 56% fat and 34% complex carbohydrate); days 2–5 provided ∼ 3150 kJ (750 kcal, 9% protein, 44% fat, and 47% complex carbohydrate). Further details of the FMD are described in the study protocol (16) and primary outcomes are reported elsewhere(17).

##### 2.2.1.2 Data collection

The time of FMD discontinuation and the reasons for discontinuation were collected through telephone calls, which took place during every FMD period. Finger prick β-ketone levels to assess the participants’ fasting state, were measured on the day directly after the 5-day cycle of the FMD in fasting condition using the FreeStyle Libre (Abbott) after the 1^st^, 4^th^, 8^th^ and 12^th^ cycle. Diabetes Treatment Satisfaction Questionnaires (DTSQ) were administered to assess treatment satisfaction; the DTSQs (status) questionnaires at baseline and after 3-, 6-, 9- and 12-month follow-up; and the DTSQc (change) questionnaire was administered at 12 months only (19, 20). The DTSQs reflects treatment satisfaction at that particular moment, whereas the DTSQc asks persons to consider whether their treatment satisfaction has changed over time. The DTSQc has six items scored on a scale from -3 to 3, with a minimum total score of -18 and a maximum score of 18. The DTSQs status has six items scored on a scale from 0 to 6 with a minimum total score of zero and a maximum score of 36. Higher scores indicate better treatment satisfaction. Both questionnaires also address hyper- and hypoglycaemia with higher scores indicating a higher frequency of having the feeling of too high or too low glucose levels.

##### 2.2.1.3 Data analyses

Data are summarized using means and standard deviations, or in the case of a skewed distribution, medians and interquartile ranges. Differences in ketone levels and DTSQs scores between the FMD group and the control group were estimated using linear mixed models. The models included fixed effects for time and time-by-arm interaction terms with random effects for individual participants. The models were adjusted for randomization stratifiers (sex and weight > 100 kg) and for the baseline value of the outcome. Intention-to-treat analyses were conducted, including all available data including data from individuals who had already discontinued the FMD diet. To compare the DTSQc (change) questionnaires at 12 months between the FMD group and the control group, an intention-to-treat analysis was conducted using independent t-tests.

### Qualitative study

#### Participants and procedure

Focus groups were chosen as they allow participants to exchange experiences, perceptions, and strategies. They were organized between December 2020 and April 2021 with participants who had completed the study year, to further explore participant’ experiences of following the FMD. Through purposive sampling, a subgroup of the FMD participants was invited to ensure diverse representations with respect to sex, age, BMI, and adherence levels. The participants provided written and verbal informed consent. During these focus groups, two themes were discussed (90- 120 minutes in total with a break between the themes) via a semi structured questionnaire (**Appendix 2**): 1) adherence to the FMD, including the barriers and facilitators involved; and 2) self-initiated lifestyle changes, and the barriers and facilitators involved in this process. The results for the first theme are described in the present paper, and the results for the second theme are described elsewhere (21).

#### Data collection

Focus groups were held online through the video platform “Jitsi” (https://meet.jit.si/) due to the measures regarding the coronavirus disease 2019 (COVID-19) outbreak. The focus groups consisted of three to four participants and were organized until data saturation was reached. While the number of participants falls below the recommended threshold for face-to-face settings (22), it was selected to ensure optimal engagement in online interactions. An experienced moderator (PP) led all the focus groups with the assistance of two other researchers (MS or EB, and CS).

##### 2.2.2.3. Data analyses

To evaluate the representativeness of our focus group participants, baseline data and the number of completed FMD cycles were compared between the whole FMD group and the focus group participants. Continuous variables were compared by using the independent t-test, or the Mann-Whitney U test if the assumption of normality was violated. Categorical outcomes were analysed with the chi-square test, or the Fisher’s exact test if the assumptions of the chi-square test were violated.

Field notes were taken and after each session observations were shared, and main themes were discussed. All focus groups were audio-recorded and transcribed verbatim by CS, and transcripts were reviewed by MS and EB. The names and other identifying data were omitted from the transcript and replaced by the study ID. Atlas.Ti version 22 for Windows was used for data analysis. Through an inductive approach, MS and CS independently analysed the text by assigning codes (strings or words/phrases) to quotes (segments of text), creating a code list. The research team (MS, CS, EB, MA and PP) discussed the codes and mapped the codes on the TDF domains and the elements of the Capability, Opportunity and Motivation Behaviour (COM-B) model(23, 24). The TDF is a widely used comprehensive, theory-informed integrated framework of the various domains describing determinants of behaviour(23). While the TDF was originally developed to analyse the behaviour of healthcare professionals(23), it is currently also used to provide a comprehensive analysis of determinants of patient behaviour(25, 26). Thereafter, by re-reading, reviewing and clustering the codes, inductive themes and sub-themes were generated to explain the data in depth (27, 28).

## Results

In the FIT trial, 129 individuals were assessed for eligibility between November 20, 2018, and July 1, 2020. One hundred persons were enrolled and randomized between the FMD group (n=51) and the control group (n=49). Eight participants were lost to follow-up before the baseline measurements were obtained (**Appendix 3**). Baseline measurements were completed for n=49 participants in the FMD group and n=43 participants in the control group (**Appendix 4**) and were comparable between groups.

### Quantitative data on adherence with the FMD

#### Discontinuation of FMD

Among the 49 participants who started the FMD intervention, 30 (61%) reported having completed all 12 cycles of the FMD programme (**Figure 1**). 19 participants (39%) discontinued the FMD diet of which fifteen (31%) discontinued because of one or more diet-related factors such as disliking the taste (n=6), experiencing adverse events including nausea, diarrhoea and fatigue (n=5), experiencing hunger (n=3), inability to combine with daily life (n=2), fear of COVID-19 by losing weight (n=1), and the unwillingness to lose any more weight (n=1). The other four participants (8%) decided to discontinue due to health issues unrelated to the FMD (n=2) or scheduling issues (n=2).

**Figure 1.**
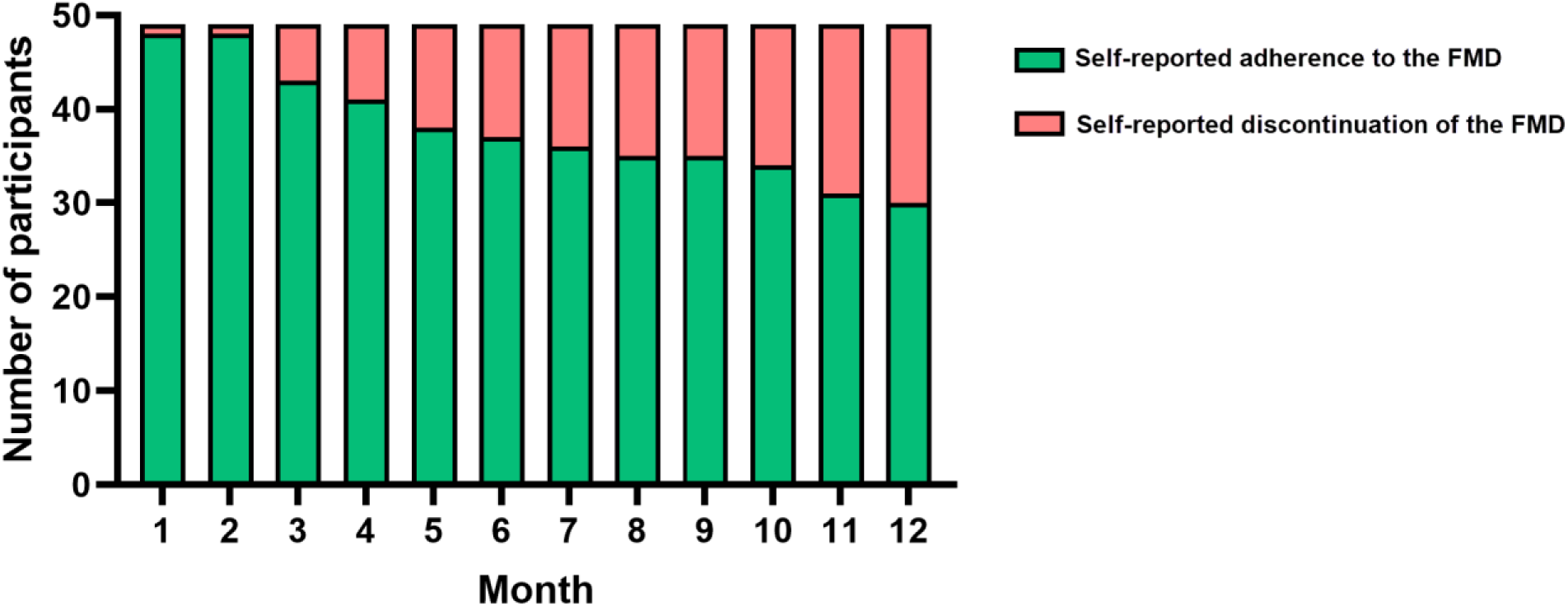
Number of participants with self-reported adherence to the FMD per month.

#### Serum ketone levels

β-Ketone levels were higher in the FMD group (which included participants who had already discontinued the FMD) than in the control group after the 1^st^, 4^th^, 8^th^ and 12^th^ cycles (**Table 1**). In the control group, β-ketone levels remained low throughout the year of follow-up.

**Table 1.** β-Ketone levels in the FMD and control groups (intention-to-treat analysis) Linear mixed models were computed with fixed effects for time and time-by-arm interaction terms and with random effects for individual participants. * The models were adjusted for the baseline value of the outcome and for randomization stratifiers (sex and weight > 100 kg). CI = confidence interval. FMD = fasting-mimicking diet. All available data were used, including ketone levels of participants who had already discontinued the FMD. Most missing data were the result of cancelled appointments during the COVID-19 period.

| Ketone levels | FMD |  | Control |  | Adjusted* Estimated treatment effect | p-value |
| --- | --- | --- | --- | --- | --- | --- |
|  | n | Mean (SD) | n | Mean (SD) |  |  |
| After 1<sup>st</sup> cycle | 40 | 0.46 (0.31) | 37 | 0.12 (0.12) | 0.36 (0.24 to 0.48) | <0.01 |
| After 4<sup>th</sup> cycle | 42 | 0.45 (0.34) | 33 | 0.19 (0.34) | 0.24 (0.11 to 0.36) | <0.01 |
| After 8<sup>th</sup> cycle | 32 | 0.43 (0.36) | 26 | 0.10 (0.04) | 0.26 (0.12 to 0.40) | <0.01 |
| After 12<sup>th</sup> cycle | 35 | 0.43 (0.40) | 28 | 0.15 (0.14) | 0.23 (0.097 to 0.36) | <0.01 |

#### Treatment satisfaction and perceived hyperglycaemia/hypoglycaemia

At the end of the trial, the participants were asked if they experienced a change in treatment satisfaction over the past year by use of the DTSQ change (DTSQc) questionnaire. There was no significant difference in DTSQ change scores between the FMD group and the control group regarding the total treatment satisfaction score, perceived hyperglycaemia or perceived hypoglycaemia (**Table 2**).

**Table 2.**
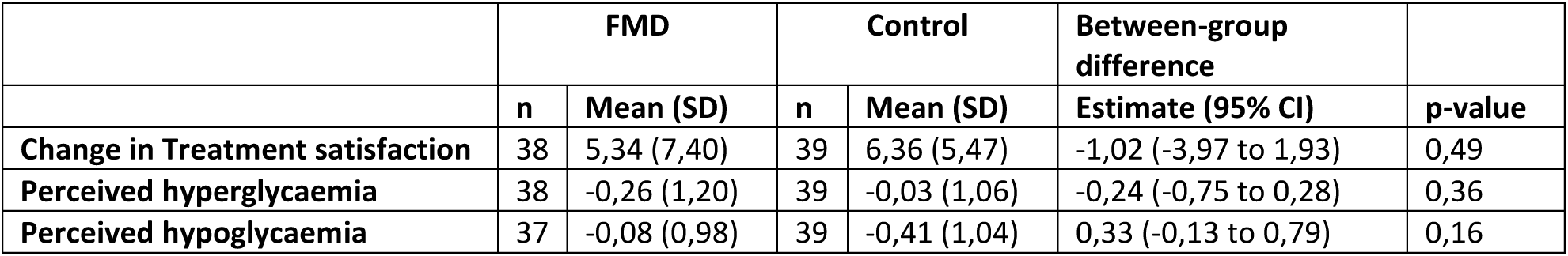
Total treatment satisfaction score and perceived hyperglycaemia and hypoglycaemia in the FMD and control group. Scores were measured using the DTSQc questionnaires via independent t-tests. All available data were used. Perceived hyperglycaemia was reported by one FMD patient. DTSQc = diabetes treatment satisfaction questionnaire (change)

Treatment satisfaction as measured by the DTSQ status (DTSQs) questionnaire, which reflects treatment satisfaction at the moment the questionnaire was administered, did not differ between the FMD group and the control group after 3, 6, 9 or 12 months compared with baseline (**Appendix 5**). Furthermore, there was no intervention effect on perceived hyperglycaemia or hypoglycaemia at these time points (**Appendix 5**).

### Barriers and facilitators of adherence to the FMD

For the focus groups, 23 persons with a diverse representation in sex, age, BMI, and adherence levels were included **(Appendix 6)**. Due to illness, work-related emergency and difficulty participating in the video call, three participants could not attend the focus groups. After six focus group sessions with 3-4 participants per group, no new facilitators or barriers emerged, and the researchers agreed that data saturation was reached. The identified facilitators and barriers were mapped onto combined COM-B and TDF model (**Figure 2**) and are described with the corresponding TDF domain presented in italics between brackets *(TDF domain)*.Facilitators and barriers could be grouped into three themes: diet-related factors, health-related factors and social interactions.

**Figure 2.**
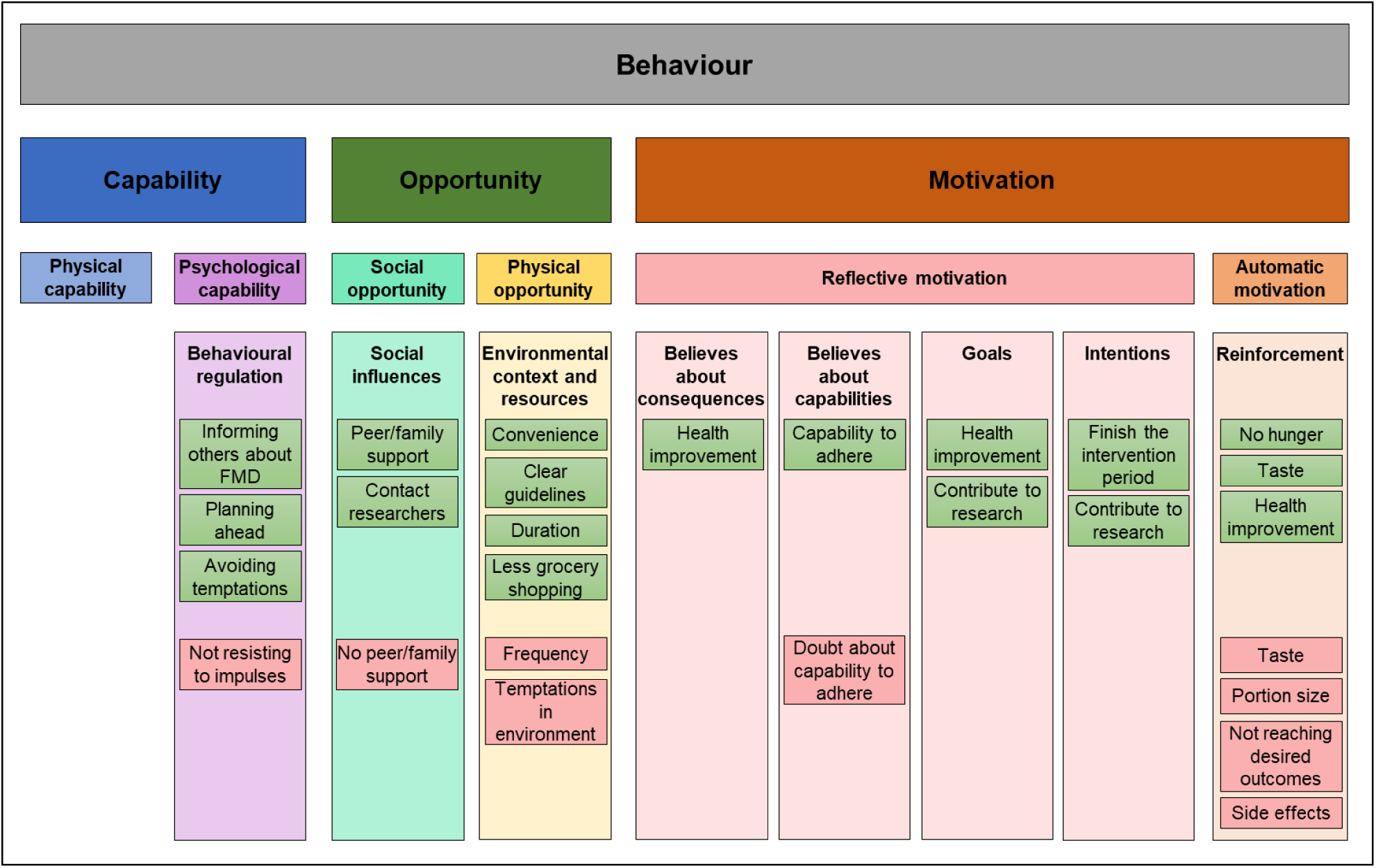
Combined TDF and COM-B models mapped with barriers (red) and facilitators (green) influencing FMD adherence. COM-B = capability, opportunity, motivation. TDF = Theoretical domains framework

#### Facilitators

##### Diet-related factors

The prepackaged meals with simple instructions were experienced as convenient and supportive *(environmental context and resources)*. The program’s clear guidelines on the specific time frames for the FMD and the permissible food options *(environmental context and resources)* were beneficial, as they were found to eliminate the need for decision-making about food consumption during FMD periods. *“The idea of doing it so many days in a row, the intermittent aspect - that appealed to me, this suits me, it gives clarity. Stop smoking, you stop smoking, and then you’re not going to grab a cigarette anymore. You can’t do that with food. But with such a box, that’s like, you stick to the diet or not- that gave me something to hold on to.” [FG1, male].* For most participants, the five consecutive days of FMD were perceived as feasible *(environmental context and resources)*. *“It’s only five days every time - and I think that’s the big difference, it’s only five days in the month. If you would do it every other week, I think it will be much harder. “[FG3, male]* A few participants even indicated that they could extend the five days or were comfortable with extending the one-year timeframe. Most participants found it easy to integrate the FMD into their work schedules. Distraction by work and the absence of food in the workplace facilitated adherence to the diet. The availability of the diet eliminated the need for grocery shopping on FMD days, reducing exposure to temptations *(environmental context and resources)*. Not feeling hungry *(reinforcement)* was often unexpected and facilitated adherence. It also worked as a facilitator when the overall taste or the taste of several components of the diet was to the participants liking *(reinforcement)* or when there was a possibility to switch products in the programme to more preferred flavours (*reinforcement*). Participants who experienced diet-related challenges adopted strategies to overcome these challenges. When feeling hungry, participants chose to go to bed early to prevent snacking. When products were disliked, alternative flavours for specific diet items were requested, or participants decided not to take the products *(behavioural regulation)*.

##### Health-related factors

Participants had the strong initial intention to finish the full year of the programme (*intentions)*. Most expected to successfully adhere to the diet (*beliefs about capabilities*). *“You leave the hospital [research unit] and then you think well I’m the man and I’m going to try it. And I have to say from the very first box I thought I could do it.” [FG1, male]* The strong internal motivation to reach desired health-related goals (*goals*) and the belief that adherence to the FMD would help them achieve these goals, acted as key facilitators (*believes about consequences*). Goals included weight loss, improved blood glucose values, reduced medication usage, and lifestyle changes. Experiencing positive health effects like weight loss, improved glucose values, reduced medication and feeling healthier in general, served as a major facilitator for most participants (*reinforcement*). *“ I stopped my diabetes medication completely, and half of my blood pressure medication as well. I learned how to eat differently, and I am doing well with my rheumatism, I can move much better, and I am really happy with it. I have much less pain.” [FG4, female]* Furthermore, contributing to research regarding T2D was a strong motivator for some participants to start and complete the FIT study year (*goals, intentions*).

##### Social interactions

Support from family members and peers (*social influences*) was crucial for adherence to the diet. Social encouragement and efforts to adjust the environment facilitated adherence. Examples include family members who adjusted their own meals, handled grocery shopping or prepared the food for the rest of the family. *“I was also motivated by my partner. At one point, my partner said she was also going to go along with my fasting diet as much as possible. - that was a lot of fun. And even after that, we tried to motivate each other a bit. Like come on, go on.” [FG1, male ].* Social activities were sometimes considered helpful, as participants took pride in sharing their achievements with others and they felt extra motivated by positive peer feedback. To manage social temptations during events, participants often planned their diet period around festivities, informed others about their diet to reduce food temptation or chose not to attend at all (*behaviour regulation*). *“I had an anniversary that was celebrated in the evening, but I thought I’m not going there because I am following the diet, and when you arrive there in the evening, and everyone walks by with tasty snacks. – I don’t want to do that to myself.” [FG3, female ]* Participants expressed positive views about their interactions with researchers during the diet period, appreciating the opportunity to share both their challenges and progress (*social influences*). *“I did experience those phone calls as pleasant, as nice, just telling them how things are going, that also motivates you and that you have some contact that you are not just a number - I liked that.” [FG6, male ].* Opinions varied on the importance of the phone calls. While some participants found them motivating and reinforcing for adhering to the FMD, others experienced them as pleasant but not crucial for adherence.

#### Barriers

##### Diet-related factors

Participants in focus groups consistently cited taste and portion size as primary barriers to adhering to the FMD diet *(reinforcement)*. The monotonic taste caused increasing resistance to the diet over time in several participants, and some struggled with small portions. “*Well, it was really very little [food]. And, uhm, I also found it monotonous” [FG1, male].* There was also longing for fresh products and some persons occasionally supplemented the FMD with other foods *(reinforcement)*. *“I managed to stick to the diet however some days I took something aside like cucumber” [FG5, female]*. The monthly frequency was sometimes mentioned as being too high or the duration of 5 days too long, negatively impacting adherence (*environmental context and resources*). Some participants would not consider continuing the programme after the first year, whereas others would only consider continuing if the frequency was reduced *(environmental context and resources)*. Another barrier was the occurrence of side effects including reduced energy, nausea, headaches, or diarrhoea. These side effects were common but generally mild (17); however for one participant, vomiting was a reason to stop (*reinforcement*). Nausea was also mentioned to occur on the first day after the fasting period when trying to eat average portion sizes or specific kinds of food. Working was sometimes assessed as difficult in combination with the diet *(environmental context and resources)*, by having less energy, or difficulties with preparing the diet at work (mainly because a microwave or a stove is needed to prepare the soups).

##### 3.3.2.2 Health-related factors

A minority initially doubted their ability to adhere to the diet, which could act as a barrier *(beliefs about capabilities)*. Disappointment arose if desired outcomes, such as improved blood glucose levels, medication reduction or weight loss, were not achieved as expected, which served as a barrier *(reinforcement)*. *“I’ve lost weight. I’ve lost eight kilos, but I expected a lot more.” [FG4, male ]*

##### 3.3.2.2 Social interactions

Adhering to the diet at home was found to be more challenging in the absence of family support and when preparing meals for the family but being unable to partake in them (*social influences*). Outside the home, participants reported additional challenges, such as discomfort in consuming diet products in the presence of others and the need to resist temptations, particularly in social settings such as parties (*behaviour regulation*).

#### Themes

All facilitators and barriers identified in the focus groups could be related to three themes; diet-related factors (including taste and quantity, structure, frequency, and side effects), health-related factors (including expectations to improve health, contribution to research for better healthcare and effects on health) and social interactions (including peer and family support, social pressure, temptations during social activities, and contact with healthcare providers). An overview of these themes is presented in **Figure 3**, which could serve as a tool for healthcare providers and their patients to identify and discuss patient-specific factors expected to influence dietary adherence.

**Figure 3.**
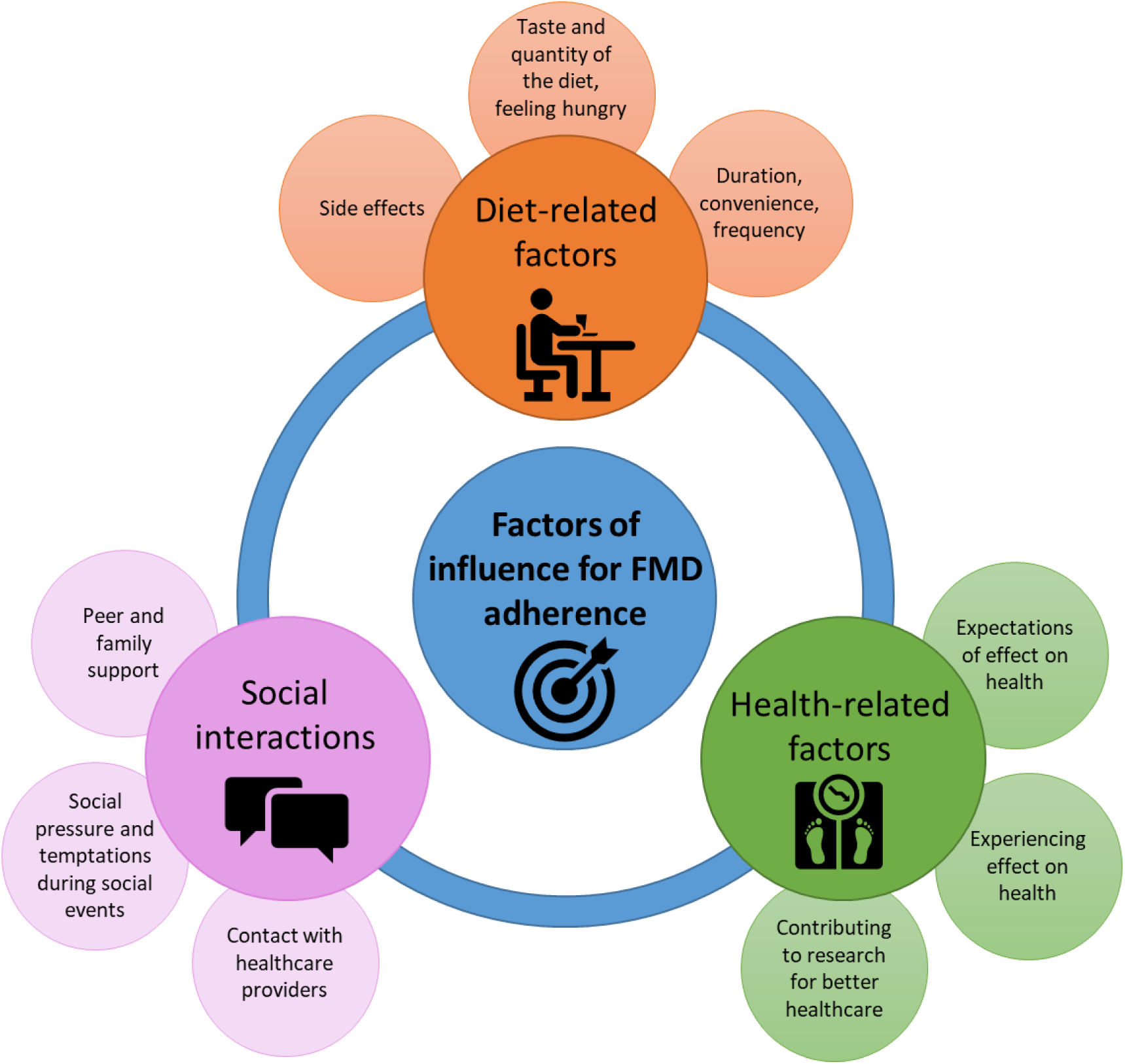
Themes influencing adherence to an FMD in persons with T2D.

## Discussion

Of the participants who started the FMD intervention, two thirds completed the one-year programme. Reasons for discontinuation of the diet included taste preferences, hunger, difficulty integrating it into daily life, concerns about weight loss during the COVID-19 pandemic, and reluctance to lose further weight. Ketone levels were consistently higher throughout the year in the FMD group than in the control group. Treatment satisfaction and perceived hypo-and hyperglycaemia did not differ between groups. Qualitative research consisting of focus groups with FMD participants revealed that convenience, short cycles, not feeling hungry, internal motivation, believing in a beneficial (health) effects, experiencing health effects and social support worked facilitating. Barriers included taste, quantity, and frequency of the FMD, environmental temptations, and lack of social support.

Adherence to dietary programs for persons with T2D has been reported in only a minority of studies. Carter et al. reported a adherence rate of 44% in an intermittent energy restriction group and 49% in a continuous energy restriction group after 12 months (29). Corley et al. found a self-reported adherence rate of 24.2% for a calorie target in an intermittent fasting programme (30). Esposito et al. reported that 24% of persons with T2D had high adherence to a Mediterranean diet (MD), scaled as 6 to 9 points on a scale of 0 to 9 of the Mediterranean Diet Score (31). Considering these reported adherence rates, the 61% adherence to the FMD programme highlights its practical feasibility.

This conclusion is further supported by the ketone levels, which are consistently higher in the FMD group than in the control group throughout the year, indicating adherence (of at least some of the participants) with the FMD programme. In comparison, results of a study with a low-carbohydrate group and a group on a conventional diet, the difference in urinary ketone levels disappeared after three months (32). Another FMD trial reported consistent blood ketone levels of 0.5 mmol/l after 3 and 6 months (15). The ketone levels in the present studies do appear to decrease over the year in the FMD group, from 0.52 mmol/l after the first cycle and 0.40 mmol/l after 12 cycles, which can be explained by the inclusion of participants of the FMD group who discontinued the FMD programme in the analyses.

The feasibility of the FMD programme is also supported by the lack of difference in treatment satisfaction. Even though participants described in the focus groups that several difficulties and side effects occurred, this did not influence treatment satisfaction scores. Furthermore, following the FMD programme did not result in greater perceived hyper-and hypoglycaemia compared to the control group.

The several facilitators and barriers to adherence that were identified in the focus groups, were either diet-related, health-related or related to social interactions. Among the diet-related factors, convenience and simplicity of the FMD emerged as key facilitators. These are valuable assets for adherence to a lifestyle intervention, as often mentioned barriers to adherence are the absence of prefabricated meal options, not fully understanding the programme and the lack of knowledge about preparing healthy food or appropriate portion sizes (33–38). The majority of the participants indicated not feeling hungry, which poses a valuable facilitator of adherence, as hunger often makes it challenging to limit portion sizes or reduce overall food consumption (34, 39). The main barrier of the FMD programme is the taste of the pre-fabricated diet, which was described as being unappealing from the start or becoming too monotonous over time. Taste has been identified as an important factor influencing adherence to other dietary interventions, including very low caloric diets (40) and meal replacement diets with severe energy restriction (41).

Health related factors such as internal motivation, goals, belief in a beneficial effect of the FMD programme on health and experiencing health changes were strong facilitators of adherence. This aligns with findings that adherence is hindered by a lack of motivation (34, 35, 39, 40), having a negative attitude toward the intervention or its guidelines (39), a lack of glycaemic control and continued disease progression despite adherence (42). Although side effects can be significant barriers in dietary interventions (40, 43), only a few participants in our study cited them as a barrier, with only one participant discontinuing the diet due to side effects.

Empirical studies have demonstrated a positive and significant relationship between social support and treatment adherence among persons with diabetes (44), which aligns with our findings.

Difficulties with social activities, such as the temptation of food during social events, were common barriers mentioned by participants and are frequently reported in the literature (33, 38, 40, 43, 45), although many participants in the current study also mentioned the strong facilitating effect of social support.

### Implications for Practice

The combination of adherence rates, ketone levels, treatment satisfaction and identified facilitators and barriers in this mixed-methods study led to the conclusion that the FMD programme was feasible for individuals with type 2 diabetes in primary care. By linking identified TDF domains of facilitators and barriers to behavioural change techniques (46, 47), several implementation recommendations have emerged that could even improve the observed high adherence to the FMD programme in primary care.

Within the *Capability* domain, spontaneous use of self-regulation strategies such as informing others about the diet, planning, or avoiding temptation by going to bed early worked as facilitators.

However, impulses were sometimes hard to resist. Healthcare providers can use motivational interviewing techniques to discuss with persons with T2D the difficulties they may encounter at home and during social events, and suggest strategies to address these difficulties such as coping planning and monitoring (48–51).

In the domain of *Opportunity*, family and peer support significantly enhanced adherence, while its absence posed a substantial barrier. It is therefore advisable to assess whether social support is present and sufficient and if there is any desire for additional support. If there is a desire for additional support, caregivers could use caregiver-hosted interventions, which have the potential to improve social support (52). The literature suggests that contact with researchers or healthcare providers can be important for participants adjusting to a new diet (33, 38), however as participants in the present study found contact nice but not necessary, healthcare providers could ask if regular contact during the FMD programme is desired. As the FMD programme had clear rules to follow, this stimulated adherence (*environmental context and resources*). The convenience, clear guidelines, duration and absence of making food choices worked facilitating, however, the frequency was often found too high. Healthcare providers could use collaborative goal-setting (48), to discuss with participants what is feasible and whether the programme should be adapted to improve adherence to the FMD.

Collaborative goal-setting could also be used by healthcare providers to support their patients in the *Motivation* domain, to discuss which goals and health improvements are realistic and feasible. Given the wide variation in health effects among participants following the FMD program, informing patients about this variability is recommended. As adherence was strongly supported by regular feedback of health improvement, these feedback moments could be facilitated by the health-care provider. In the reinforcement domain, adherence might be improved by informing patients about the possible side effects and how to manage them. Furthermore, since taste was a significant barrier to adherence, offering the option to substitute disliked products with comparable alternatives that better suit individual preferences could improve adherence.

### Strengths and Limitations

The strength of the current study lies in its combination of objective adherence measurements and qualitative research on facilitators of and barriers to FMD adherence. This comprehensive approach allows persons with T2D and healthcare providers to make informed decisions about FMD suitability for individuals, support persons with T2D during the program, and optimize adherence. However, there are limitations. The FIT trial was not designed or powered to detect changes in ketone levels and DTSQ scores. As the participants in this study also indicated that research goals were facilitating, this might have enhanced adherence to the FMD. The high FMD adherence rate (80%) among focus group participants compared with the overall FMD group (61%) may affect representativeness.

Participants with positive health effects might have been more enthusiastic to share their experiences during the focus groups than participants who did not complete the 12 FMD cycles. However, valuable feedback on barriers was gathered from both compliant and noncompliant participants.

### Conclusion

Considering the number of participants who successfully adhered to the one-year periodic FMD program, the measured ketone levels, the treatment satisfaction and the facilitators and barriers expressed by interviewed FMD participants, a periodic FMD programme appears feasible for many persons with T2D in primary care compared to other lifestyle-intervention programs. Additionally, adherence could be enhanced by addressing the various facilitators and barriers identified, including the taste of the FMD, the positive effect on health status and the importance of social support.

## Acknowledgements

## Data Availability

The datasets used during the current study are available upon reasonable request. Requests for access to data should be sent to the FIT trial corresponding email. All proposals requesting data access will need to specify how the data will be used, and all proposals will need approval of the trial co-investigator team before data release.

## Acknowledgements

We gratefully acknowledge the contributions of all the participants, the trial steering committee, the general practice centres, the supporting staff and the research nurses involved in the FIT trial.

## Author contributions

Marjolein Schoonakker: Conceptualisation, Methodology, Project administration, Investigation, Formal analysis, Writing- Original draft preparation, Visualization. Elske van den Burg: Conceptualisation, Methodology, Project administration, Investigation, Writing – Review & Editing. Carlijn Sturm: Methodology, Investigation, Formal analysis. Hanno Pijl: Funding acquisition, Conceptualisation, Methodology, Writing – Review & Editing, Supervision. Mattijs Numans: Funding acquisition, Conceptualisation, Methodology, Writing – Review & Editing, Supervision. Hildo Lamb: Funding acquisition, Conceptualisation, Methodology, Writing – Review & Editing, Supervision.

Marieke Adriaanse: Methodology, Writing – Review & Editing, Supervision. Petra van Peet: Conceptualisation, Methodology, Investigation, Writing – Review & Editing, Supervision.

## Abbreviations

BMI =: Body Mass Index.
CI =: confidence interval
COM-B =: Capability Opportunity and Motivation Behaviour model.
COVID-19 =: coronavirus disease 2019.
DTSQs =: Diabetes Treatment Satisfaction Questionnaires Status
DTSQc =: Diabetes Treatment Satisfaction Questionnaire Change
FIT trial =: Fasting In diabetes Treatment trial.
FMD =: fasting-mimicking diet.
HbA1c =: glycated haemoglobin.
LUMC =: Leiden University Medical Centre.
N =: number.
SD =: standard deviation.
T2D =: type 2 diabetes.
TDF =: Theoretical Domains Framework.

## References

1. VZinfo.nl (2022): https://www.vzinfo.nl/diabetes-mellitus, RIVM: Bilthoven, 10-05-2023. [

2. International Diabetes Federation, IDF diabetes atlas, 10th edition. 2021. https://diabetesatlas.org/atlas/tenth-edition/ (accessed Mar 15, 2022). .

3. Lean MEJ, Leslie WS, Barnes AC, Brosnahan N, Thom G, McCombie L, et al. Primary care-led weight management for remission of type 2 diabetes (DiRECT): an open-label, cluster-randomised trial. Lancet. 2018;391(10120):541–51.

4. Lean MEJ, Leslie WS, Barnes AC, Brosnahan N, Thom G, McCombie L, et al. Durability of a primary care-led weight-management intervention for remission of type 2 diabetes: 2-year results of the DiRECT open-label, cluster-randomised trial. The lancet Diabetes & endocrinology. 2019;7(5):344–55.

5. McKenzie AL, Hallberg SJ, Creighton BC, Volk BM, Link TM, Abner MK, et al. A Novel Intervention Including Individualized Nutritional Recommendations Reduces Hemoglobin A1c Level, Medication Use, and Weight in Type 2 Diabetes. JMIR Diabetes. 2017;2(1):e5.

6. Kelly J, Karlsen M, Steinke G. Type 2 Diabetes Remission and Lifestyle Medicine: A Position Statement From the American College of Lifestyle Medicine. Am J Lifestyle Med. 2020;14(4):406–19.

7. Li S, Du Y, Meireles C, Sharma K, Qi L, Castillo A, et al. Adherence to ketogenic diet in lifestyle interventions in adults with overweight or obesity and type 2 diabetes: a scoping review. Nutr Diabetes. 2023;13(1):16.

8. Petroni ML, Brodosi L, Marchignoli F, Sasdelli AS, Caraceni P, Marchesini G, et al. Nutrition in Patients with Type 2 Diabetes: Present Knowledge and Remaining Challenges. Nutrients. 2021;13(8).

9. MacDonald CS, Ried-Larsen M, Soleimani J, Alsawas M, Lieberman DE, Ismail AS, et al. A systematic review of adherence to physical activity interventions in individuals with type 2 diabetes. Diabetes Metab Res Rev. 2021;37(8):e3444.

10. van den Burg EL, van Peet PG, Schoonakker MP, van de Haar DE, Numans ME, Pijl H. Metabolic impact of intermittent energy restriction and periodic fasting in patients with type 2 diabetes: a systematic review. Nutrition Reviews. 2023.

11. Grajower MM, Horne BD. Clinical Management of Intermittent Fasting in Patients with Diabetes Mellitus. Nutrients. 2019;11(4).

12. Horne BD, Grajower MM, Anderson JL. Limited Evidence for the Health Effects and Safety of Intermittent Fasting Among Patients With Type 2 Diabetes. Jama. 2020;324(4):341–2.

13. Wei M, Brandhorst S, Shelehchi M, Mirzaei H, Cheng CW, Budniak J, et al. Fasting-mimicking diet and markers/risk factors for aging, diabetes, cancer, and cardiovascular disease. Science translational medicine. 2017;9(377).

14. Tang F, Lin X. Effects of Fasting-Mimicking Diet and Specific Meal Replacement Foods on Blood Glucose Control in Patients with Type 2 Diabetes: A Randomized Controlled Trial. Oxid Med Cell Longev. 2020;2020:6615295.

15. Sulaj A, Kopf S, von Rauchhaupt E, Kliemank E, Brune M, Kender Z, et al. Six-Month Periodic Fasting in Patients With Type 2 Diabetes and Diabetic Nephropathy: A Proof-of-Concept Study. J Clin Endocrinol Metab. 2022;107(8):2167–81.

16. van den Burg EL, Schoonakker MP, van Peet PG, van den Akker-van Marle ME, Willems van Dijk K, Longo VD, et al. Fasting in diabetes treatment (FIT) trial: study protocol for a randomised, controlled, assessor-blinded intervention trial on the effects of intermittent use of a fasting-mimicking diet in patients with type 2 diabetes. BMC Endocr Disord. 2020;20(1):94.

17. van den Burg EL, Schoonakker MP, van Peet PG, van den Akker-van Marle EM, Lamb HJ, Longo VD, et al. Integration of a fasting-mimicking diet programme in primary care for type 2 diabetes reduces the need for medication and improves glycaemic control: a 12-month randomised controlled trial. Diabetologia. 2024.

18. Edmonds WA, Kennedy TD. An Applied Guide to Research Designs: Quantitative, Qualitative, and Mixed Methods. 2017 2024/03/31. Thousand Oaks Thousand Oaks, California: SAGE Publications, Inc. Second. Available from: https://methods.sagepub.com/book/an-applied-guide-to-research-designs-2e.

19. Bradley C. Diabetes treatment satisfaction questionnaire. Change version for use alongside status version provides appropriate solution where ceiling effects occur. Diabetes Care. 1999;22(3):530–2.

20. Bradley C, Gamsu DS. Guidelines for encouraging psychological well-being: report of a Working Group of the World Health Organization Regional Office for Europe and International Diabetes Federation European Region St Vincent Declaration Action Programme for Diabetes. Diabet Med. 1994;11(5):510–6.

21. van den Burg EL, Schoonakker MP, Korpershoek B, Sommeling LE, Sturm CA, Lamb HJ, et al. Self-initiated lifestyle changes during a fasting-mimicking diet programme in patients with type 2 diabetes: a mixed-methods study. BMC Primary Care. 2024;25(1):148.

22. Carlsen B, Glenton C. What about N? A methodological study of sample-size reporting in focus group studies. BMC Medical Research Methodology. 2011;11(1):26.

23. Atkins L, Francis J, Islam R, O’Connor D, Patey A, Ivers N, et al. A guide to using the Theoretical Domains Framework of behaviour change to investigate implementation problems. Implement Sci. 2017;12(1):77.

24. Michie S, van Stralen MM, West R. The behaviour change wheel: a new method for characterising and designing behaviour change interventions. Implement Sci. 2011;6:42.

25. Flannery C, McHugh S, Anaba AE, Clifford E, O’Riordan M, Kenny LC, et al. Enablers and barriers to physical activity in overweight and obese pregnant women: an analysis informed by the theoretical domains framework and COM-B model. BMC Pregnancy Childbirth. 2018;18(1):178.

26. Quayyum F, Dombrowski SU. Barriers to nutritional pregnancy preparation and support needs in women and men: Qualitative study based on the Theoretical Domains Framework. Womens Health (Lond). 2021;17:17455065211042182.

27. Huberman A, Miles M, Ritchie J, Spencer L. Qualitative Data Analysis for Applied Policy Research.

28. Braun V, Clarke V. Using thematic analysis in psychology. Qualitative Research in Psychology. 2006;3:77–101.

29. Carter S, Clifton PM, Keogh JB. Effect of Intermittent Compared With Continuous Energy Restricted Diet on Glycemic Control in Patients With Type 2 Diabetes: A Randomized Noninferiority Trial. JAMA Network Open. 2018;1(3):e180756-e.

30. Corley BT, Carroll RW, Hall RM, Weatherall M, Parry-Strong A, Krebs JD. Intermittent fasting in Type 2 diabetes mellitus and the risk of hypoglycaemia: a randomized controlled trial. Diabet Med. 2018;35(5):588–94.

31. Esposito K, Maiorino MI, Di Palo C, Giugliano D. Adherence to a Mediterranean diet and glycaemic control in Type 2 diabetes mellitus. Diabet Med. 2009;26(9):900–7.

32. Foster GD, Wyatt HR, Hill JO, McGuckin BG, Brill C, Mohammed BS, et al. A randomized trial of a low-carbohydrate diet for obesity. N Engl J Med. 2003;348(21):2082–90.

33. Mathew R, Gucciardi E, De Melo M, Barata P. Self-management experiences among men and women with type 2 diabetes mellitus: a qualitative analysis. BMC Fam Pract. 2012;13:122.

34. Laranjo L, Neves AL, Costa A, Ribeiro RT, Couto L, Sá AB. Facilitators, barriers and expectations in the self-management of type 2 diabetes--a qualitative study from Portugal. Eur J Gen Pract. 2015;21(2):103–10.

35. Booth AO, Lowis C, Dean M, Hunter SJ, McKinley MC. Diet and physical activity in the self-management of type 2 diabetes: barriers and facilitators identified by patients and health professionals. Primary Health Care Research & Development. 2013;14(3):293–306.

36. Wilson D, Diji AK, Marfo R, Amoh P, Duodu PA, Akyirem S, et al. Dietary adherence among persons with type 2 diabetes: A concurrent mixed methods study. PLoS One. 2024;19(5):e0302914.

37. Tripathi D, Vikram NK, Chaturvedi S, Bhatia N. Barriers and facilitators in dietary and physical activity management of type 2 diabetes: Perspective of healthcare providers and patients. Diabetes & Metabolic Syndrome: Clinical Research & Reviews. 2023;17(3):102741.

38. Lee SA, Sypniewski C, Bensadon BA, McLaren C, Donahoo WT, Sibille KT, et al. Determinants of Adherence in Time-Restricted Feeding in Older Adults: Lessons from a Pilot Study. Nutrients. 2020;12(3).

39. Deslippe AL, Soanes A, Bouchaud CC, Beckenstein H, Slim M, Plourde H, et al. Barriers and facilitators to diet, physical activity and lifestyle behavior intervention adherence: a qualitative systematic review of the literature. Int J Behav Nutr Phys Act. 2023;20(1):14.

40. Roesler A, Marshall S, Rahimi-Ardabili H, Duve E, Abbott K, Blumfield M, et al. Choosing and following a very low calorie diet program in Australia: A quasi-mixed methods study to understand experiences, barriers, and facilitators in a self-initiated environment. Nutr Diet. 2021;78(2):202–17.

41. Maston G, Franklin J, Hocking S, Swinbourne J, Gibson A, Manson E, et al. Dietary adherence and program attrition during a severely energy-restricted diet among people with complex class III obesity: A qualitative exploration. PLoS One. 2021;16(6):e0253127.

42. Nagelkerk J, Reick K, Meengs L. Perceived barriers and effective strategies to diabetes self-management. J Adv Nurs. 2006;54(2):151–8.

43. Lawford BJ, Bennell KL, Jones SE, Keating C, Brown C, Hinman RS. "It’s the single best thing I’ve done in the last 10 years": a qualitative study exploring patient and dietitian experiences with, and perceptions of, a multi-component dietary weight loss program for knee osteoarthritis. Osteoarthritis Cartilage. 2021;29(4):507–17.

44. Miller TA, Dimatteo MR. Importance of family/social support and impact on adherence to diabetic therapy. Diabetes Metab Syndr Obes. 2013;6:421–6.

45. Parr EB, Devlin BL, Radford BE, Hawley JA. A Delayed Morning and Earlier Evening Time-Restricted Feeding Protocol for Improving Glycemic Control and Dietary Adherence in Men with Overweight/Obesity: A Randomized Controlled Trial. Nutrients. 2020;12(2).

46. Michie S, Wood CE, Johnston M, Abraham C, Francis JJ, Hardeman W. Behaviour change techniques: the development and evaluation of a taxonomic method for reporting and describing behaviour change interventions (a suite of five studies involving consensus methods, randomised controlled trials and analysis of qualitative data). Health Technol Assess. 2015;19(99):1–188.

47. Michie S, Richardson M, Johnston M, Abraham C, Francis J, Hardeman W, et al. The behavior change technique taxonomy (v1) of 93 hierarchically clustered techniques: building an international consensus for the reporting of behavior change interventions. Ann Behav Med. 2013;46(1):81–95.

48. Salvia MG, Quatromoni PA. Behavioral approaches to nutrition and eating patterns for managing type 2 diabetes: A review. Am J Med Open. 2023;9:100034.

49. Spahn JM, Reeves RS, Keim KS, Laquatra I, Kellogg M, Jortberg B, et al. State of the evidence regarding behavior change theories and strategies in nutrition counseling to facilitate health and food behavior change. J Am Diet Assoc. 2010;110(6):879–91.

50. Sniehotta F, Schwarzer R, Scholz U, Schüz B. Action planning and coping planning for long-term lifestyle change: Theory and assessment. European Journal of Social Psychology. 2005;35:565–76.

51. Braun M, Schroé H, De Paepe AL, Crombez G. Building on Existing Classifications of Behavior Change Techniques to Classify Planned Coping Strategies: Physical Activity Diary Study. JMIR Form Res. 2023;7:e50573.

52. Clayton C, Motley C, Sakakibara B. Enhancing Social Support Among People with Cardiovascular Disease: a Systematic Scoping Review. Curr Cardiol Rep. 2019;21(10):123.

