## Appendix for "Feasibility of following a fasting-mimicking diet programme in persons with type 2 diabetes – a mixed-methods study"

#### Supplementary information

### Appendix 1

#### Example meal plan of the fasting-mimicking diet for study participants in the FIT trial

| | Day 1 | Day 2 | Day 3 | Day 4 | Day 5 |
|---|---|---|---|---|---|
| **Breakfast** | Tea<br><br>Nut bar<br>Algal Oil capsule | Tea<br><br>Nut bar | Tea<br><br>Nut bar | Tea<br><br>Nut bar | Tea<br><br>Nut bar<br>Algal Oil capsule |
| **Lunch** | Tomato Soup<br>Olives<br>Kale crackers<br>Vitamin capsule | Tea<br>Mushroom Soup<br>Olives<br><br>Vitamin capsule | Tea<br><br>Tomato Soup<br>Kale Crackers<br><br>Vitamin capsule | Tea<br>Vegetable Soup<br>Olives<br><br>Vitamin capsule | Tea<br><br>Tomato Soup<br>Kale Crackers<br><br>Vitamin capsule |
| **Afternoon** | Tea<br>Nut bar | Tea<br>Olives | Tea | Tea<br>Olives | Tea |
| **Dinner** | Minestrone Soup<br>Choco crisp bar<br>Vitamin capsule | Tea<br>Quinoa Mix Soup<br><br>Choco crisp bar<br><br>Vitamin capsule | Tea<br>Minestrone Soup<br><br>Vitamin capsule | Tea<br>Quinoa Mix Soup<br><br>Choco crisp bar<br><br>Vitamin capsule | Tea<br>Minestrone Soup<br><br>Vitamin capsule |
| **During the day** | | Syrup for water flavouring | Syrup for water flavouring | Syrup for water flavouring | Syrup for water flavouring |

### Appendix 2

#### Semi-structured questionnaire for focus group discussions in the FIT trial

**Fasting In diabetes Treatment**
Experiences with following a fasting-mimicking diet: qualitative research

**Introductory round** *(Analysed in the current study)*

During the study, you participated in the diet.
- How did you feel about the diet before the trial?
- What did you hope to achieve by following the diet?
- How many cycles of the diet did you complete?
- When did you complete the follow-up of the FIT study?

**Theme 1: Compliance with the fasting-mimicking diet** *(analysed in the current study)*

**General questions**
- Can you tell me about your experience with the diet?
- Before starting the study, did you expect to complete the 12 dietary cycles?

**Specific questions**

1. Barriers and facilitators
- What made it difficult for you to adhere to the diet?
- What made it easier for you to adhere to the diet?
- Did the COVID-19 period affect your ability to maintain the diet?

2. Possible role of primary healthcare professionals
- What could help you to maintain the diet?
- Is there a potential role for primary healthcare professionals?

*Possible topics to address if the discussion stalls*

**Possible factors that could play a role in whether or not the diet is maintained:**
- Form of the diet (a ready-made box)
- Short duration (five days per month)
- Taste
- Side effects
- Fixed dates for the diet weeks
- Telephone appointments during the diet week
- Outpatient appointment on day 6 of the diet week
- Influence of weight change
- Influence of change in laboratory results (HbA1c, glucose)
- Impact of other health problems
- Environmental factors
    - Role of partner/spouse
    - Role of friends/family

**-- Break --**

**Theme 2: Impact of following the fasting-mimicking diet on lifestyle** (analysed elsewhere)

**General questions**

- Can you tell us something about your lifestyle during the year in which you participated in the FIT trial?
- Did following the diet affect your lifestyle?
- Were you motivated to change your lifestyle when you started the FIT trial?
- Did you plan to make other lifestyle changes during the study and did you feel you would succeed in doing so?
- What made it more difficult for you to change your lifestyle?
- What made it easier for you to change your lifestyle?

**Specific questions**

1. Influence on dietary pattern

- Did your usual eating pattern change during the time you regularly followed the diet?
- Did following the diet make you think differently about nutrition?
- Was there a period of "overeating" / binge eating / more snacking between diet periods?

2. Influence on exercise pattern

- Did your normal exercise pattern change during the time you regularly followed the diet?
- Did following the diet change your thoughts about exercise?
- Did you have enough energy to keep moving during the dietary cycles?

3. Possible role of healthcare professionals

- What might help to modify your usual lifestyle while following the diet intermittently?
- Is there possibly a role for primary care professionals?

4. Lifestyle during the corona period

- Did the COVID-19 period affect your lifestyle?

**-- End --**

### Appendix 3

#### Flow chart of participant inclusion in the FIT trial

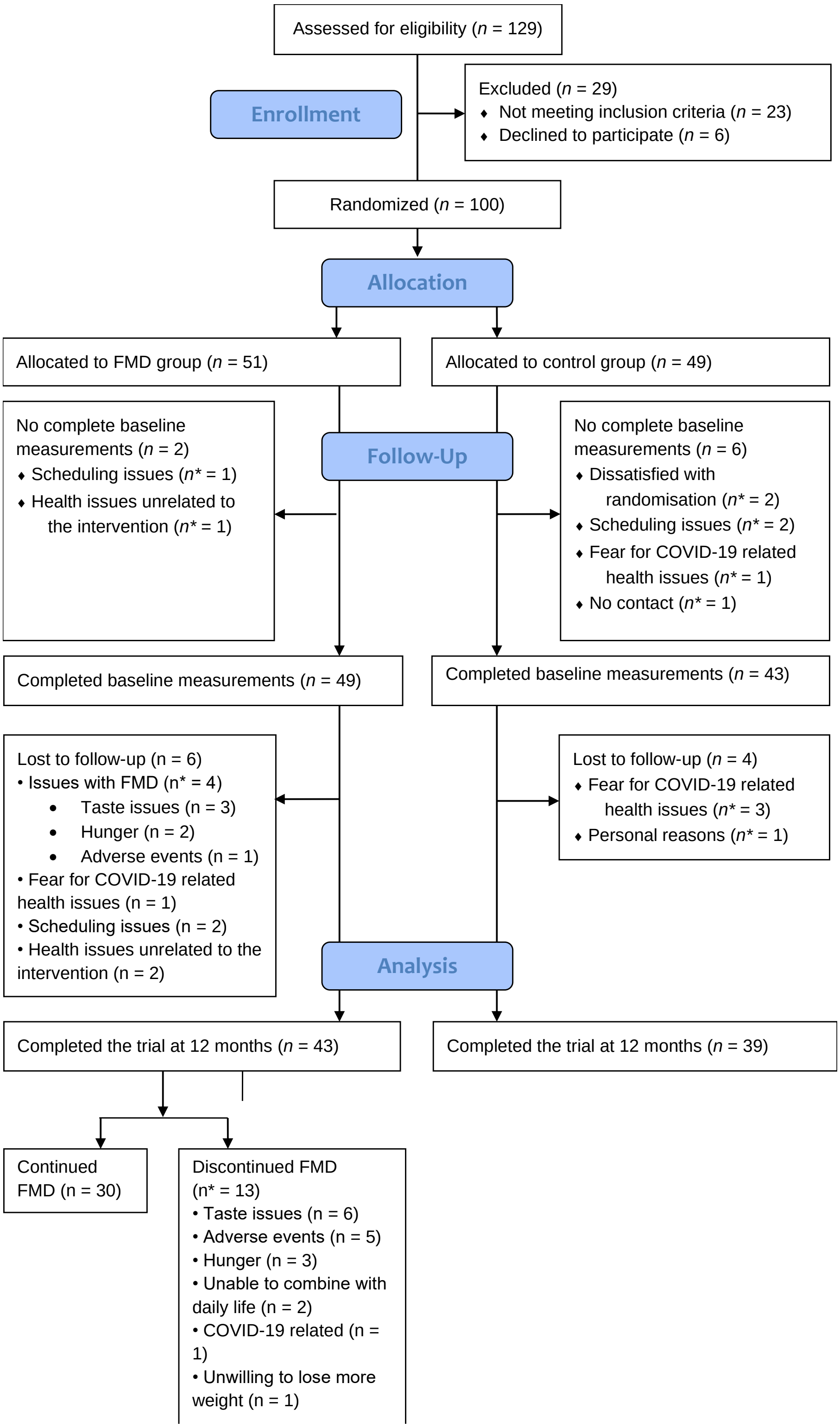

COVID-19 = coronavirus disease 2019. FMD = fasting-mimicking diet. n = number of participants.
n* = number of participants for whom this was reason for being lost to follow-up or discontinuing FMD, there may be several reasons per participant.

### Appendix 4

#### Demographics and baseline characteristics (n=92)

| | **FMD group (*n* = 49)** | **Control group (*n* = 43)** |
|---|---|---|
| **Demographics** | | |
| Age (years), mean ± SD | 62 ± 8 | 64 ± 8 |
| Sex, *n* (%) | | |
| Male | 26 (53) | 22 (51) |
| Female | 23 (47) | 21 (49) |
| **Medical History** | | |
| Time since diagnosis T2D (years), median (IQR) | 4 (3-12) | 6 (3-10) |
| Use of metformin, *n* (%) | 46 (94) | 36 (84) |
| **Laboratory measurements** | | |
| HbA1c (%), mean ± SD | 6.9 ± 0.87 | 7.1 ± 1.13 |
| HbA1c (mmol/mol), mean ± SD | 52.2 ± 9.3 | 53.7 ± 12.2 |
| **Anthropometrics** | | |
| BMI (kg/m$^2$), median (IQR) | 33.2 ± 4.9 | 32.7 ± 3.7 |
| **DTSCs** | | |
| Treatment satisfaction | 30.2 ± 5.6 | 30.3 ± 4.3 |
| Perceived frequency of hyperglycaemia | 1.3 ± 1.7 | 1.4 ± 1.9 |
| Perceived frequency of hypoglycaemia | 0.8 ± 1.5 | 0.7 ± 1.2 |

Data are presented as mean ± SD, median (IQR) or number (*n*) with percentage (%). BMI=Body Mass Index. DTSQs=diabetes treatment satisfaction questionnaire (status). FMD=fasting-mimicking diet. HbA1c=glycated haemoglobin. T2D=type 2 diabetes.

### Appendix 5

#### Changes in Treatment satisfaction, hyperglycaemia and hypoglycaemias with Adjusted Treatment Effects of FMD vs control group

| | | FMD | | Control | | Adjusted* Estimated treatment effect (95% CI) | p-value |
|---|---|---|---|---|---|---|---|
| | | **n** | **mean (sd)** | **n** | **mean (sd)** | | |
| **Treatment satisfaction** | Baseline | 50 | 30.3 (5.64) | 43 | 30.3 (4.30) | | |
| | 3 months | 46 | 31.1 (4.99) | 38 | 30.7 (6.12) | 0.54 (-1.4 to 2.5) | 0.59 |
| | 6 months | 45 | 30.0 (5.23) | 36 | 31.3 (4.39) | -1.4 (-3.4 to 0.58) | 0.17 |
| | 9 months | 43 | 30.5 (5.97) | 38 | 30.1 (5.86) | 0.47 (-1.5 to 2.4) | 0.64 |
| | 12 months | 41 | 28.0 (6.39) | 40 | 29.4 (5.54) | -1.7 (-3.7 to 0.26) | 0.094 |
| **Hyperglycaemia** | Baseline | 50 | 1.32 (1.70) | 43 | 1.40 (1.94) | | |
| | 3 months | 48 | 0.90 (1.49) | 41 | 0.76 (1.61) | 0.065(-0.49 to 0.62) | 0.82 |
| | 6 months | 46 | 0.91 (1.23) | 38 | 0.90 (1.66) | 0.053 (-0.52 to 0.62) | 0.86 |
| | 9 months | 42 | 1.05 (1.56) | 39 | 1.33 (1.78) | -0.32(-0.90 to 0.26) | 0.29 |
| | 12 months | 41 | 0.85 (1.53) | 39 | 1.41 (1.96) | -0.45 (-1.0 to 0.13) | 0.13 |
| **Hypoglycaemia** | Baseline | 50 | 0.74 (1.47) | 43 | 0.67 (1.19) | | |
| | 3 months | 48 | 0.81 (1.28) | 41 | 0.49 (1.00) | 0.29 (-0.15 to 0.74) | 0.20 |
| | 6 months | 46 | 0.59 (0.93) | 38 | 0.50 (1.06) | 0.11 (-0.35 to 0.56) | 0.65 |
| | 9 months | 43 | 0.51 (0.10) | 39 | 0.95 (1.50) | -0.42 (-0.88 to 0.037) | 0.075 |
| | 12 months | 41 | 0.76 (1.28) | 40 | 0.38 (0.90) | 0.46 (-0.0029 to 0.92) | 0.054 |

Scores were measured using the DTSQs questionnaires.
The linear mixed models were computed with fixed effects for time and time-by-arm interaction terms and with random effects for individual participants.
* The models were adjusted for the baseline value of the outcome and for randomization stratifiers (sex and weight > 100 kg).
CI = confidence interval. FMD = fasting-mimicking diet.

### Appendix 6

#### Characteristics of all FMD participants (n = 49) compared to the FMD participants in the focus groups (n=20)

| | **FMD group (*n* = 49)** | **Focus group participants (n=20)** |
|---|---|---|
| **Demographics** | | |
| Age (years), mean ± SD | 62 ± 8 | 63 ± 8 |
| Sex, *n* (%) | | |
| Male | 26 (53) | 10 (50) |
| Female | 23 (47) | 10 (50) |
| **Medical History** | | |
| Time since diagnosis T2D (years), median (IQR) | 4 (3-12) | 5 (2-10) |
| Use of metformin, *n* (%) | 46 (94) | 18 (90) |
| **Laboratory measurements** | | |
| HbA1c (%), mean ± SD | 7.1 ± 1.13 | 6.8 ± 2.9 |
| HbA1c (mmol/mol), mean ± SD | 52.2 ± 9.3 | 50.8 ± 8.1 |
| **12 month compliance, *n* (%)** | 30 (61) | 16 (80) |

Data are presented as mean ± SD, median (IQR) or number (*n*) with percentage (%).
BMI = Body Mass Index. FMD = fasting-mimicking diet. HbA1c = glycated haemoglobin. IQR = interquartile range. n = number. SD = standard deviation. T2D = type 2 diabetes.
